# Transcriptomic and Metabolomic analyses in Monozygotic and Dizygotic twins

**DOI:** 10.1101/2024.06.25.24309452

**Authors:** Nikki Hubers, Gabin Drouard, Rick Jansen, René Pool, Jouke Jan Hottenga, Miina Ollikainen, Xiaoling Wang, Gonneke Willemsen, Jaakko Kaprio, Dorret I. Boomsma, Jenny van Dongen

**Affiliations:** Department of Biological Psychology, Vrije Universiteit Amsterdam, Amsterdam, the Netherlands; Amsterdam Reproduction & Development (AR&D) research institute, Amsterdam, the Netherlands; Amsterdam Public Health research institute, Amsterdam, the Netherlands; Institute for Molecular Medicine Finland (FIMM), HiLIFE, University of Helsinki, Helsinki, Finland; Amsterdam UMC location Vrije Universiteit Amsterdam, Department of Psychiatry & Amsterdam Neuroscience - Complex Trait Genetics (VUmc) and Mood, Anxiety, Psychosis, Stress & Sleep; Minerva Foundation Institute for Medical Research, Helsinki, Finland; Georgia Prevention Institute, Medical College of Georgia, Augusta University, Augusta, GA, USA; Faculty of Health, Sport and Wellbeing, Inholland University of Applied Sciences, Haarlem, the Netherlands; Department of Complex Trait Genetics, Center for Neurogenomics and Cognitive Research, Amsterdam, Vrije Universiteit Amsterdam

**Keywords:** Twinning, Twins, Transcriptomics, Metabolomics, Multi-omics

## Abstract

Monozygotic (MZ) and dizygotic (DZ) twins are studied to understand genetic and environmental influences on complex traits, however the mechanisms behind twinning are not completely understood. (Epi)genomic studies identified SNPs associated with DZ twinning and DNA methylation sites with MZ twinning. To find molecular biomarkers of twinning, we compared transcriptomics and metabolomics data from MZ and DZ twins. We analyzed 42,663 RNA transcripts in 1,453 MZ twins and 1,294 DZ twins from the Netherlands Twin Register (NTR), followed by sex-stratified analyses. The top 5% transcripts with lowest p-values were analyzed for replication in 217 MZ and 158 DZ twins from the older Finnish Twin cohort (FTC). In the NTR, one transcript (*PURG)* was significantly differentially expressed between MZ and DZ twins; but this did not replicate in FTC. Pathway analyses highlighted the WNT-pathway, previously associated with MZ twinning, and the TGF-B and SMAD pathway, previously associated with DZ twinning. Meta-analysis of 169 serum metabolites in 2,797 MZ and 2,040 DZ twins from the NTR, FTC and FinnTwin12, showed no metabolomic differences. Overall, we did not find replicable transcript-level expression differences in blood between MZ and DZ twins, but highlighted the TGF-B/SMAD pathway as a potential transcriptional biomarker for DZ twinning.

## Introduction

Twinning is defined as the process that gives rise to dizygotic (DZ) or monozygotic (MZ) twins, and in rarer occasions to triplets and other higher-order multiples (van Dongen et al., 2023). MZ and DZ twins are often studied to provide insight into the genetic and environmental influences on complex traits through the classical twin design or other twin-family designs (Hagenbeek et al., 2023; Visscher, 2004). However, much remains unclear about the etiology of MZ and DZ twinning. DZ twinning is the result of a spontaneous double ovulation, runs in families and can be studied as a model for heightened fertility (Beck et al., 2021; Hall, 2003). MZ twins arise after a fertilized egg cell splits, but the mechanisms behind this are still largely a mystery (Hall, 2021b; van Dongen et al., 2021, 2023).

Genome-wide association studies (GWAS) of DZ twinning highlighted multiple genes such as *FSHB, FSHR, SMAD3* and *GNRH,* with obvious roles in female reproduction (Mbarek et al., 2016, 2024). An epigenome-wide association study (EWAS) highlighted DNA methylation differences at over 800 sites in blood from MZ twins when compared to DZ twins and singletons (van Dongen et al., 2021). The methylation differences in blood also replicated in buccal cells. Still, mechanisms leading to the formation of twins, in particular MZ twins, are not fully understood (Hall, 2021a). Furthermore, it is unknown whether molecular markers of twinning might be found in other omics layers that have not yet been studied in connection to twinning, including the transcriptome and metabolome. Since differentially methylated positions (DMPs) in MZ twins are strongly enriched in polycomb-repressed regions and heterochromatin (van Dongen et al 2021), which are typically associated with transcriptionally-silenced developmental genes and other transcriptionally repressed genes in somatic tissues, MZ-DMPs are a priori expected not to be accompanied by transcriptional differences in somatic tissues. Nevertheless, this hypothesis remains to be tested. For DZ twinning, no epigenetic or transcriptomic signatures have been identified to date. Studying these layers may provide additional insights into the etiology and can lead to the identification of biomarkers for MZ and DZ twinning (Johnson et al., 2016; Soininen et al., 2015; Whipp et al., 2022).

Transcriptomics is the study of all RNA molecules from protein coding to noncoding RNA (Thompson et al., 2016). Data from protein coding RNAs are sometimes integrated with GWAS results to identify novel gene-trait associations (Yin et al., 2022). These so-called transcriptome-wide association studies (TWASs) are usually performed as post-GWAS analyses using databases such as GTEx and tools like SMR/HEIDI (Gamazon et al., 2015; Zhu et al., 2016). In the largest DZ twinning GWAS, eight genes were identified using a TWAS analysis in several female fertility related tissues: *ARL14EP, CAPRIN2, ZFPM1, SMAD3, MPPED2-AS, GOLGA8T, PCBP2,* and *FAM66D*. Data from twins have also been studied to estimate the heritability of RNA sequence levels (Ouwens et al., 2019), but RNA transcripts have not been analyzed to investigate twinning etiologies.

Metabolomics is the study of the small molecules involved in cellular metabolism (Patti et al., 2012). Unlike other omics, metabolomics provides tools to measure biochemical activity directly, by monitoring the substrates and products involved in cellular metabolism. Studies focusing on metabolomics are widely performed in search of biomarkers for physical and mental health conditions, but have never been performed before for twinning (Guijas et al., 2018).

In this study, we compare serum metabolomics and blood transcriptomics profiles between MZ and DZ twins to enhance the discovery of molecular biomarkers of twinning. We include adult participants from the Netherlands Twin Register (NTR) and from two Finnish twin cohorts, FinnTwin12 (FT12) and the older Finnish Twin Cohort (FTC) of whom many have been involved in one of the two previous omics studies into twinning (Kaprio et al., 2019; Ligthart et al., 2019; Mbarek et al., 2024; Rose et al., 2019; van Dongen et al., 2021). For the transcriptomics, we analyzed gene expression levels of 42,663 RNA transcripts in 2,747 participants of the NTR and repeated our analysis in the 5% transcripts with the lowest p-values in 375 participants of the FTC. For the metabolomics, the three cohorts had measurements from the same platform allowing us to meta-analyze 169 metabolite levels from 4,837 DZ and MZ twins.

## Methods

### Participants

#### NTR

The NTR is a population based cohort that has been collecting data from twins and their families since the 1980’s (Ligthart et al., 2019). In 2004 the NTR started a large-scale biological sample collection of nearly 10,000 participants to create a resource for future omics and biomarker studies (Willemsen et al., 2010). During a home visit in the morning, eight tubes of fasting blood and a morning urine sample were collected along with phenotypic information on health, medication use, body composition and smoking. For fertile women samples were obtained, as much as possible, on days 3–5 of their menstrual cycle or in the pill-free week if on oral contraception. RNA data were generated with the Affymetrix U219 array for 3,362 participants of the NTR, of whom 2,828 were twins. From these twins we excluded 81 individuals who were pregnant or did not have complete covariate data leading to a total of 2,747 twins who we included in our study (Table 1). Metabolomics data were collected for 4,227 participants of the NTR of whom 3,638 were twins with complete phenotypic information that could be included in this study. Zygosity of the participants was determined using genotype information, blood group information or multiple survey items (Ligthart et al., 2019).

**Table 1:** The descriptives of the participants included in the study. NTR = Netherlands Twin register; FT12 = FinnTwin12; FTC = the older Finnish twin cohort. A: the current smokers included the occasional smokers.

|  | **Metabolomics** |  |  | **Transcriptomics** |  |
| --- | --- | --- | --- | --- | --- |
|  | **NTR** | **FTC** | **FT12** | **NTR** | **FTC** |
| **Participants** | 3,638 | 435 | 764 | 2,747 | 375 |
| **MZ twins (%)** | 2,117 (58.2%) | 250 (55.8%) | 430 (44.7%) | 1,453 (52.9%) | 217 (59.1%) |
| **Mean age at time of biological sample (sd)** | 36.1 (11.1) | 62 (4.4) | 22.3 (1.7) | 38.3 (11.6) | 62.3 (3.8) |
| **N female (%)** | 2,465 (67.8%) | 259 (59.5%) | 473 (57.2%) | 1,797 (65.4%) | 222 (59.2%) |
| **Mean BMI (sd)** | 24 (3.8) | 27.3 (4.9) | 23.3 (3.8) | 24.0 (3.9) | 27.3 (4.7) |
| **N Fasting yes (%)** | 3,087 (85.8%) | 435 (100%) | 764 (100%) | 2,623 (95.5%) | 375 (100%) |
| **N Lipid Medication (%)** | 82 (2.3%) | 0 (0%) | 0 (0%) | - | - |
| **N current smoking<sup>a</sup> (%)** | 772 (21.4%) | 73 (16.8%) | 334 (44.0%) | 667 (24.3%) | 58 (15.5%) |

#### FTC

One subset of the Finnish Twin Cohort is the older Finnish Twin Cohort (FTC), which includes twins born before 1958 (Wright et al., 2014). Four waves of questionnaires have been sent to unselected members of the cohort in 1975, 1981, 1990 and 2011, respectively (Kaprio et al., 2019). In 2015, a selected subset of these twins came in for measurement of their blood pressure, completed interviews and questionnaires and provided a fasting blood sample for biochemical measures, and samples for omics (Huang et al., 2018). Metabolomic data for 435 participants were included in the study. Out of the 402 peripheral blood samples, high quality RNA was obtained for 391 subjects and 375 twins of them had complete phenotypic data and could be included in the current study (Table 1). Zygosity of the twins was determined by the first questionnaires for twins and confirmed by genotyping.

#### FT12

FT12 is a population-based longitudinal cohort including twins born in Finland between 1983 and 1987 (Rose et al., 2019). FT12 was designed for baseline assessments at an early age preceding onset of regular exposure to alcohol, tobacco or other substances (Rose et al., 2019). Subsequent waves of follow-up in FT12 at ages 11-12, 14, 17, and 22 years, have created a rich dataset of behavioral assessments from multiple sources across three stages of adolescence and into early adulthood. The “age 22” assessment wave involved 1,347 twin individuals, with 779 individuals attending in-person assessments and thus venous blood plasma samples could be collected (Table 1; (Whipp et al., 2022)). We removed 15 participants from the data due to pregnancy or lipid lowering medication, resulting in a total sample of 764 participants from FT12. Zygosity was determined using questionnaires at baseline and confirmed later by genotyping. No transcriptomics data were available in the FT12.

### Omics data

#### Transcriptomics

##### NTR

The generation of the gene expression arrays and data in the NTR was described before (Jansen et al., 2014; Wright et al., 2014). Briefly, peripheral venous blood samples were drawn in the morning (7:00—11:00 a.m.) after an overnight fast. Within 20 minutes of sampling, heparinized whole blood was transferred into PAXgene Blood RNA tubes (Qiagen) and stored at -20°C. The frozen PAXgene tubes were shipped to the Rutgers University and DNA Repository (RUCDR, http://www.rucdr.org). RNA was extracted by the Qiagen Universal liquid handling system, as per the manufacturer’s protocol. RNA quality and quantity was assessed by Caliper AMS90 with HT DNA5K/RNA LabChips. Samples were hybridized to Affymetrix U219 array plates (GeneTitan) which contains 530,467 probes for 49,293 transcripts. Array hybridization, washing, staining, and scanning were carried out in an Affymetrix GeneTitan System per the manufacturer’s protocol. Twin pairs were randomized over the sample plates. Expression data were required to pass standard Affymetrix Expression Console quality metrics before further undergoing quality control. Samples were excluded if they showed sex inconsistency. Log2 transformation and quantile normalization were applied to the gene expression data. In the main analyses we excluded 1,578 transcripts located on the X and Y chromosome, leading to a total of 42,663 RNA transcripts included in main analysis, 44,182 in the female-only analysis and 44,241 in the male-only analysis.

##### FTC

An in-depth description of the transcriptomics measurements of the transcriptomics data collection can be found in Huang et al (2018) (Huang et al., 2018). RNA samples were extracted from the peripheral leukocytes stored in the RNA cell protection reagents (QIAGEN Inc. Valencia, CA, USA) using the miRNeasy Mini Kit (QIAGEN Inc. Valencia, CA, USA). Transcriptome-wide gene expression data were obtained using the Illumina HumanHT-12 v4 Expression BeadChip (Illumina Inc) which targets more than 48,000 transcripts that provide transcriptome-wide coverage of well-characterized genes, gene candidates and splice variants. A block design was used to keep the distributions of sex and zygosity similar across chips (12 samples/chip) with the co-twins assigned to the same chip. The twin pairs were randomly assigned as pairs to the 12 positions on each chip. Quantification was performed employing the Genome-Studio Gene Expression Module and the lumi R package for data preprocessing and quality control. The quality controls consisted of two key steps: 1; Transcripts with detection P-value <0.05 in more than 50% of the samples were defined as “present”; 2; Log2 transformation and quantile normalization were applied to the gene expression data. There were 19,530 transcripts that passed the quality control steps and they were used as indices of gene expression levels in further analyses. For the replication analyses, we selected the top 2,133 (5% lowest p-value fraction) RNA transcripts from the analyses in NTR and selected the same transcript in the FTC data. We selected the probes based on genetic location and/or gene name. A total of 1,325 transcripts shared overlap between the two platforms and were included in the replication analyses.

The analysis of the transcriptomics data of the FTC cohort showed a high inflation (lambda = 1.3) of the p-values (Supplementary figure 1). This is potentially due to the decision to retain MZ and DZ twin pairs on the same RNA chips, as we observe a high correlation of 0.75 between the RNA chips and age (Supplementary Figure 2), which can introduce multi-collinearity. To address this, we repeated our analyses of zygosity in 1,325 randomly selected RNA (excluding the initial 1,325 probes) transcripts in the FTC data. When repeating the analyses to random RNA transcriptomics data, we do observe several transcripts surpassing our significance threshold after multiple testing correction burden, but when looking at the enrichment analyses we do not observe any pathways associated with MZ or DZ twinning before (Supplementary Figures 3-4).

#### Metabolomics

In all three cohorts, metabolites were measured on the metabolomics platform from Nightingale (Soininen et al., 2015). The panel included amino acids (alanine, glutamine, histidine, isoleucine, leucine, phenylalanine, tyrosine, and valine) and ketone bodies (acetate, acetoacetate, and 3-hydroxybutyrate), lipids and fatty acids. All metabolite data were available in units mmol/L, nm, g/L or μmol/L (Supplementary Table 1). The metabolomics data were pre-processed for each of the three cohorts similarly, but separately.

In the NTR, the metabolomics data were collected in different batches, which were all pre-processed separately (Hagenbeek et al., 2020; Pool et al., 2020). Metabolites were excluded from the analysis when the mean coefficient of variation exceeded 25% and the missing rate exceeded 5%. Metabolite measurements were set to missing if they were below the lower limit of detection or quantification or could be classified as an outlier (five standard deviations greater or smaller than the mean). If missing, the metabolite was imputed with half of the value of this limit, or when this limit was unknown with half of the lowest observed level for this metabolite.

In the FT12 and FTC, the metabolomics data were measured in one batch each. Metabolite values below the limit of detection were reported as missing, as were metabolite values that deviated from the mean by more than 5 standard deviations (SD). Metabolites with more than 10% missing values were discarded. Missing values were imputed in the FT12 and using the sample minimum of each metabolite for which imputation was to be performed. Outlier presence was assessed by verifying that no participant had at least one of its first three principal components greater than or less than 5 SD from the mean.

In all three cohorts, metabolites were normalized by inverse normal rank transformation so that the distribution of metabolites are of mean 0 and variance 1 (Demirkan et al., 2015; Kettunen et al., 2016). In FT12, 119 metabolites could be included and in NTR and FTC all 169 metabolites were included (Supplementary Table 1 & 2).

### Statistical analyses

#### Transcriptomics

We performed logistic regression for each of the 42,663 RNA transcripts in the NTR twin status (MZ twins coded as = 1 and DZ twins coded as= 2) as a predictor. All analyses were performed in R version 4.3.0. The models were run by using Generalized estimating equations (GEE) to account for family structure (package = *geepack_1.3.11*; function = *geelm).* We included sex, biological age at blood sampling, body mass index (BMI) at time of blood sampling, plate, well and cell counts of blood cells including neutrophiles, lymphocytes, monocytes, eosinophils and basophiles, as covariates. Given that transcriptomics data are correlated (Jansen et al., 2014), adjusting p-values to correct for multiple testing was based on the number of independent dimensions in the data. We performed principal component analyses of the transcripts (Jollife & Cadima, 2016), and corrected the transcriptomic analyses of twins for the number of effective tests performed. 2,814 principal components captured 95% of the total variation in the RNA transcript levels (Supplementary Table 3), leading to a correction where significance is reached if a p-value is below 0.05/2,814 = 1.78E-05. In addition, we ran sex stratified analyses without the sex covariate and we performed sensitivity analyses of the female-only analyses in 669 females after excluding all participants taking contraceptives and post-menopausal women (N=1,128). In the analyses of the females we additionally included the 1,519 transcripts on the X chromosome and in the analyses of the males we included the X chromosome transcripts and 59 transcripts of the Y chromosome.

We took forward the 5% fraction of transcripts with the lowest p-value in NTR in the main analyses (2,213 transcripts in 1,876 genes) and searched for matching transcripts in the FTC data. We identified 1,325 transcripts from the Illumina chip for replication, based on the location of the transcripts and the gene in which the transcript is located and included in both platforms. Given that these transcripts have already been preselected, a more stringent Bonferroni correction was used for replication in the FTC, leading to a p-value to be lower than 0.05/1,325=3.77E-05 to be significant.

#### Metabolomics

We performed logistic regression for each metabolite with twin zygosity as a predictor (MZ and DZ twin, coded 1 and 2), for each cohort separately. The covariates for each cohort comprise age, sex, smoking status, BMI, batch, and additionally only for NTR fasting status (yes/no) and use of lipid lowering medication. All variables were measured at time of blood sampling. All analyses were performed in R version 4.3.0. The models were run by using GEE to account for family structure (package = *geepack _1.3.11*; function = *geelm*). For the meta-analyses of NTR, FT12 and FTC, we applied fixed effect meta-analyses (package = *metafor4.6-0*; function = *rma*). We estimated the number of effective tests using the method of Matrix Spectral Decomposition (Chen et al., 2022). In total, we included 169 metabolites which resulted in a total of 27 effective tests, leading to a p-value to be lower than 0.05/27 = 1.85E-03 to be significant, based on Bonferroni correction.

### Enrichments analyses

#### Enrichr

We performed enrichment analysis using Enrichr on the 5% most significant transcripts in NTR and the 19 genes of the 21 significant transcripts in FTC. Enrichr is a gene set search engine that enables the querying of hundreds of thousands of annotated gene sets (Xie et al., 2021). Enrichr uniquely integrates knowledge from many high-profile projects to provide synthesized information about mammalian genes and gene sets. We employed the *enrichR_3.2* R package as an R interface to the Enrichr database (Kuleshov et al., 2016). We used the databases Gene Ontology 2023 molecular functions database (GO2023) and KEGG pathway database (KEGG) to look at the enrichment (Consortium et al., 2023; Kanehisa et al., 2023).

#### BIOS consortium database

We performed a look-up of the 5% most significant transcripts in the NTR data and the significantly different expressed transcripts in FTC in the methylation–expression association (expression quantitative trait methylation; eQTM) database of the BIOS consortium (Bonder et al., 2017; Zhernakova et al., 2016). The eQTM database did not include participants from NTR. The database contains information on associations between DNA methylation levels and gene expression, based on RNA sequencing data from ∼2000 whole blood samples. We checked whether the identified RNA transcripts identified in the current study were located in genes that showed overlap with the eQTMs of the CpGs associated with MZ twinning from our earlier EWAS study (van Dongen et al., 2021).

## Results

### Transcriptomics

We compared 42,663 RNA transcripts between 1,453 MZ and 1,291 DZ twins of the NTR (Table 1). One RNA transcript, located at 11727318_at (chr8:30853321:30854213), was significantly differentially expressed between MZ and DZ twins (higher expression in MZ twins; beta = 1.78E-02; p value = 6.19E-06). The transcript maps to chromosome 8 located in the *PURG* gene (Figure 1; Supplementary Figure 5; Supplementary table 4). We performed enrichment analyses in the GO2023 and the KEGG dataset on the genes in which 5% transcripts with the lowest p-values are located (2,133 transcripts in 1,876; Figure 2; Supplementary table 5-6). In the GO2023 database, 56 pathways were significantly enriched in the genes and the most strongly enriched pathway was ”molecular function of the estrogen response element binding”. The KEGG database highlighted 24 significant enrichment of pathways involved in breast cancer and cell adhesion elements, as well as the WNT-signaling pathway. We performed a look-up of the 5% most significant transcripts in the BIOS consortium eQTM database and identified that two transcripts (located in the genes *HTT* and *FANC)* that were significantly associated with DNA methylation of CpG sites in the genes previously found to be differentially methylated in MZ twins (P<3e-08; Supplementary table 7). Three methylation sites in the *HTT* gene (cg01807241, cg02566259 and cg06421590) showed lower methylation in MZ twins while in our result the *HTT* is higher expressed in the MZ twins. In line with this, in the BIOS dataset, lower methylation correlated with higher expression. The methylation site in *FANCC*, cg14127626, also showed less methylation in the BIOS dataset and more gene expression in the MZ twins.

**Figure 1:**
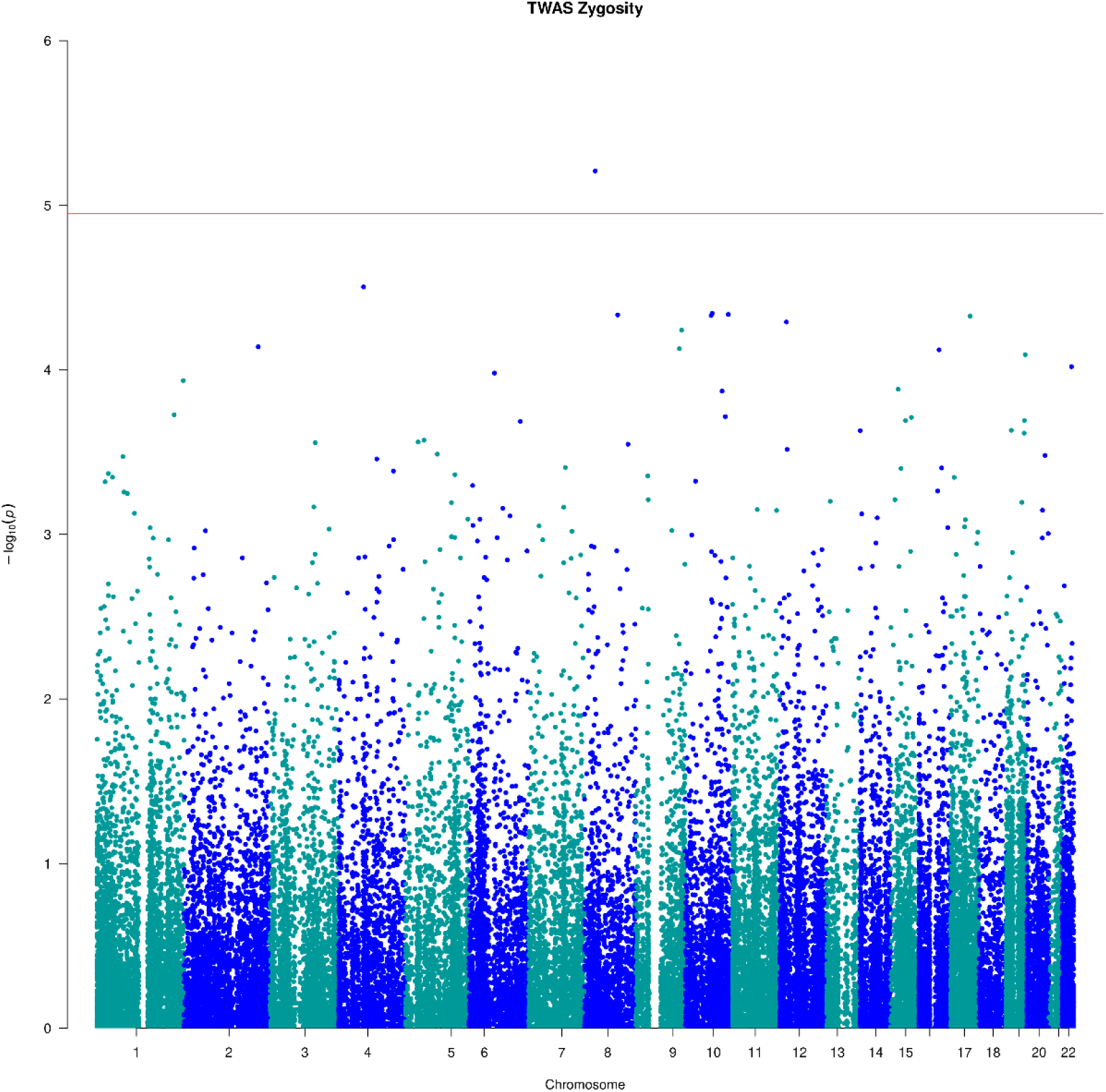
Manhattan plot showing the results of the transcriptomics analyses in the Netherlands Twin Register (NTR) comparing monozygotic and dizygotic twins.

**Figure 2:**
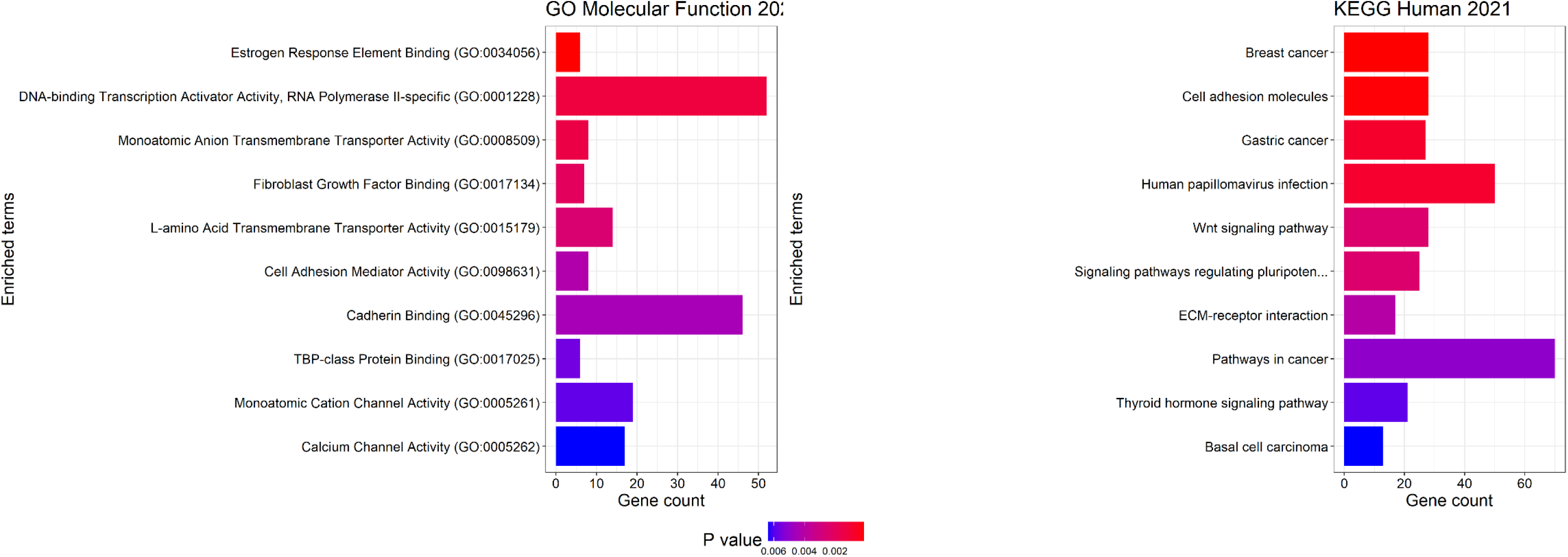
Enrichment analyses of the 5% transcripts with lowest p-values RNA transcript with the lowest p-value in the NTR. The figure only shows the ten most enriched pathways and molecular functions. The left figure shows the enrichment in the Gene Ontology 2023 molecular functions database and the right figure shows the enrichment in the KEGG human pathway database.

### Sex-stratified analyses

We reran the analyses in females and males separately, including the sex chromosomes. In the female-only analyses we included 1,008 MZ twins and 789 DZ twins and 44,182 RNA transcripts. In these analyses the same transcript in *PURG*, 11727318_at (chr8:30853321:30854213), showed significantly higher expression in MZ twins (beta = 2.50E-02; p value = 2.65E-07). In addition, we found four more transcripts, 11745032_a_at (ch4:114821440:114821440), 11736249_x_at (chr15:64657193:64658274), 11732371_at (chr19:51479729:51480947) and 11759996_at (chr5:35960858:35963053) which significantly associated with zygosity in the female-only analyses (Supplementary table 8). These transcripts are of the genes *ARSJ* (beta = -2.30E-02; p value = 1.04E-05), *KIAAA0101* (beta = -3.19E-02; p value = 1.26E-05), *KLK7* (beta = -2.43E-02; p value = 1.42E-05) and *UGT3A1* (beta = -3.81E-02; p value = 1.73E-05), respectively. We further performed enrichment analyses in the GO2023 and the KEGG dataset on the genes in which the 5% transcripts with the lowest p-value are located (2,009 transcripts; Figure 3; Supplementary tables 9-10). In total 44 pathways were enriched in the KEGG dataset and 58 function in GO2023. The strongest enriched pathways were the DNA binding Transcription Activator Activity, RNA Polymerase II-specific and pathways in cancer.

**Figure 3:**
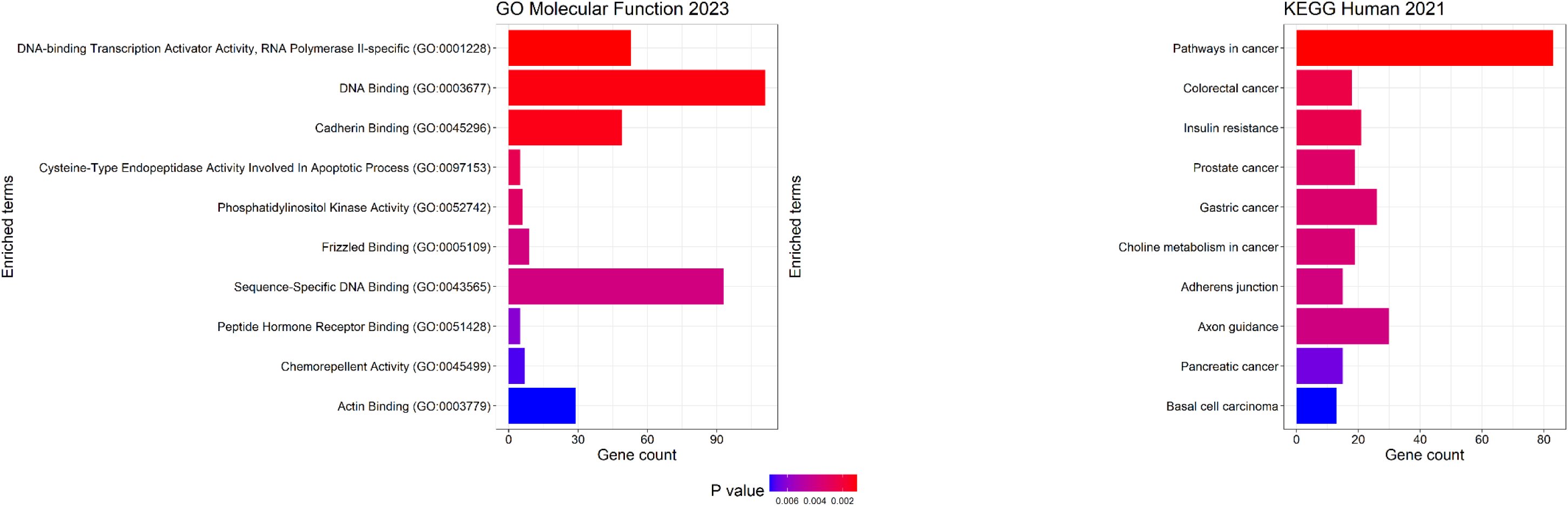
Enrichment analyses of the 5% transcripts with lowest p-values RNA transcript with the lowest p-value in the NTR females. The figure shows the ten most enriched pathways and molecular functions. The left figure shows the enrichment in the Gene Ontology 2023 molecular functions database and the right figure shows the enrichment in the KEGG human pathway database.

We performed a sensitivity analysis in the NTR female-only sample after removing all females that took contraceptives or indicated to be past menopause (N=1,128) at the time of the biological sampling (new N=669) In this analysis, six genes in the SMAD pathway (*SMAD2*, *SMAD4*, *TGFB1*, *SMAD5*, *TGFBR1*, *SMAD7*), previously indicated for DZ twinning, were downregulated in the female DZ twins based on a nominal significance (p<0.05). In addition, the R-SMAD pathway was the most significantly enriched (Supplementary tables 16-19), which was also among the enriched pathways in the female-only analyses (Supplementary table 10).

In the male-only analyses we included 445 MZ twins and 505 DZ twins and 44,241 RNA transcripts, including both sex chromosomes. In this analysis the transcript in *PURG* was not significantly different in MZ and DZ twins (beta = 7.00E-03; p value = 0.26). However, we observed three other transcripts, 11739295_a_at (chr21:45651163: 45655445), 11738638_a_at (chr20:2397878:2413399) and 11726545_x_at (chr12: 100550135: 100550836), that were significantly associated with zygosity in the male-only analyses (Supplementary table 11). These transcripts are located respectively in the *ICOSLG* (beta = 3.93E-02; p value = 4.08E-07), *TGM6* (beta = 3.22E-02; p value = 7.29E-06) and *GOLGA2P5* (beta = 3.43E-02; p value = 1.50E-05) genes. We further performed enrichment analyses in the GO2023 and the KEGG dataset on the genes in which the 5% most significant transcripts are located (2,112 transcripts; Figure 4; Supplementary Tables 12-13). In total 21 pathways were enriched in the KEGG dataset and 55 functions in GO2023. The most strongly enriched pathways were the Ankyrin Binding pathway and the Thyroid hormone signaling pathway.

**Figure 4:**
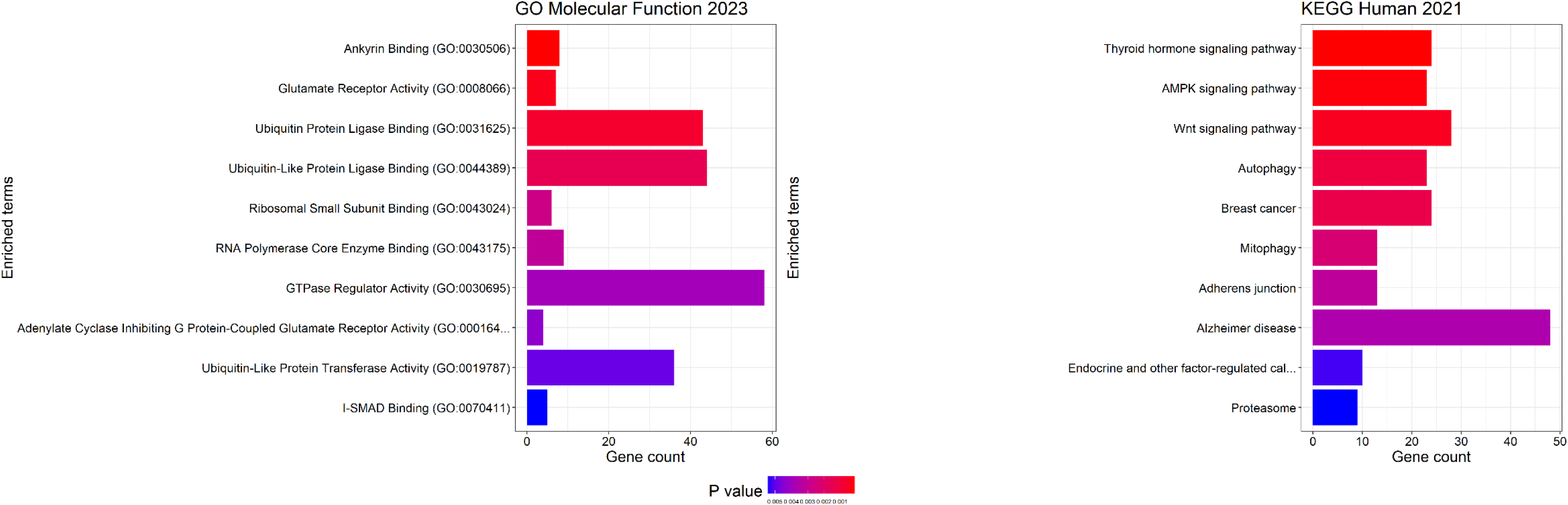
Enrichment analyses of the 5% transcripts with lowest p-values RNA transcript with the lowest p-value in the NTR males. The figure shows the ten most enriched pathways and molecular functions. The left figure shows the enrichment in the Gene Ontology 2023 molecular functions database and the right figure shows the enrichment in the KEGG human pathway database.

### Replication analyses

Replication analyses were carried out for 1,325 transcripts of the 2,133 (5%) most significant ones in the results in NTR in 375 twins of the FTC cohort (Table 1). The 1,325 transcripts show an overall correlation of -0.097 (Supplement Figure 6). We observed that 21 transcripts, from 19 genes, were differentially expressed in the FTC (Supplementary table 14). Eight of these genes showed lower expression levels in the MZ twins while the other 11 showed higher expression in the MZ twins. We performed enrichment analyses in the GO2023 and KEGG databases on the 19 genes (Figure 5). The GO2023 indicated 10 significant enrichments including the cAMP regulatory function and the R-SMAD pathway (Supplementary table 15). The TGF-B pathway was also enriched in the KEGG database, together with the cell cycle, the WNT-pathway and 7 other pathways (Supplementary table 16). A look-up of the 19 genes of the 21 significant transcripts in the BIOS consortium eQTM database did not reveal associations with CpGs that were previously associated with MZ twinning. The NTR findings for *PURG* were not replicated. For the FTC cohort the most significant transcript (ILMN_7746) lies in the *JUN* gene (beta = -1.165; p value < 1E-10). Of the 19 genes, 7 (37%) showed a replication of the same direction of effect in the NTR (Table 2).

**Figure 5:**
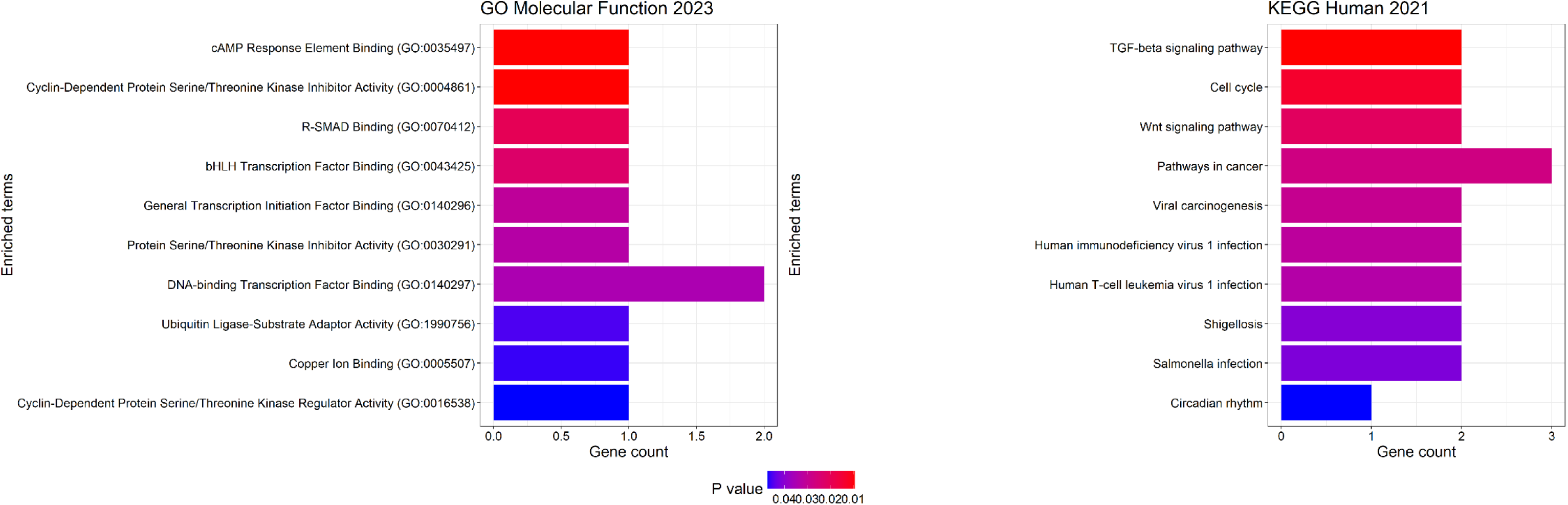
Enrichment analyses of the 19 genes from the 21 significant (p<3.77E-05) RNA transcripts for the transcriptomics analyses in the FTC. The figure shows the ten most enriched pathways and molecular functions. The left figure shows the enrichment in the Gene Ontology 2023 molecular functions database and the right figure shows the enrichment in the KEGG human pathway database.

**Table 2:** The nominal significant probes in the replication analysis in the older Finnish twin cohort (FTC) with their corresponding information for the Netherlands twin register (NTR); SE: standard error; CHR: chromosome

| Gene | Probeset id<br>Affimetrix | Probeset id<br>Illumina | CHR | Beta<br>NTR | SE<br>NTR | P-value<br>NTR | Beta<br>FTC | SE<br>FTC | P value<br>FTC |
| --- | --- | --- | --- | --- | --- | --- | --- | --- | --- |
| *ZNF564* | 11727148_s_at | ILMN_1977 | 19 | 0.014 | 0.006 | 0.012 | 0.067 | 0.013 | 2.266E-08 |
| *LEPROTL1* | 11716264_at | ILMN_4515 | 8 | 0.011 | 0.004 | 0.010 | 0.160 | 0.032 | 2.902E-07 |
| *CD69* | 11748146_at | ILMN_167129 | 12 | -0.011 | 0.005 | 0.015 | -0.486 | 0.090 | 4.691E-07 |
| *USP46* | 11725800_a_at | ILMN_364882 | 4 | -0.011 | 0.004 | 0.010 | -0.081 | 0.017 | 8.547E-07 |
| *ANKRD46* | 11761048_at | ILMN_9031 | 8 | 0.009 | 0.003 | 0.002 | 0.081 | 0.016 | 1.283E-06 |
| *CCS* | 11720676_s_at | ILMN_176820 | 11 | 0.006 | 0.003 | 0.039 | 0.045 | 0.010 | 9.794E-06 |
| *CCS* | 11720676_s_at | ILMN_23509 | 11 | 0.006 | 0.003 | 0.039 | 0.072 | 0.015 | 1.057E-05 |
| *ANKRD13C* | 11732313_at | ILMN_30045 | 1 | -0.009 | 0.005 | 0.034 | -0.043 | 0.011 | 3.654E-05 |

### Metabolomics

We meta-analyzed 169 metabolites in 2,797 MZ twins and 2,040 DZ from the NTR, FT12 and the FTC (Table 1) with a fixed effect model. The results of the individual analyses of each cohort and the meta-analyses are summarized in supplementary table 2. The results show that none of the metabolite levels differed significantly between the MZ and DZ twins after multiple testing correction (alpha=2.96E-04). Five metabolites showed differences between the MZ and DZ twins, on the nominal p-value of 0.05. The levels of glycoprotein acetyls, linoleic acid, omega-6 fatty acids, albumin and phospholipids in medium LDL were lower in the MZ twins. In three out of these five metabolites the effects of the NTR and the FT12 show the same direction, while the FTC shows the opposite effect (Supplementary figure 7). We found that the overall correlations in the betas between the three cohorts were weak with a range from -0.29 to 0.24 (Supplementary Figure 8).

## Discussion

In this study we compared transcriptomic and metabolomic data between MZ and DZ twins of the NTR, FT12 and the FTC. We show differences in the RNA transcript levels between MZ and DZ twins in genes whose relations to twinning have partially been established through other, previous, omics studies (GWAS, and EWAS) and show that the differences between the transcriptome in MZ and DZ twins depend on sex. We find limited overlap in the differentially expressed gene transcripts with the MZ twinning epigenetic signature. The enrichment analyses hint at some pathways that were detected in both the NTR and the FTC, namely, the WNT-pathways and the TGF-B pathway, that have been indicated before with either MZ or DZ twinning (Mbarek et al., 2024; van Dongen et al., 2021).

We found one transcript located in the *PURG* gene to be significantly associated with zygosity in the total NTR sample, but this finding did not replicate in FTC. When dividing the NTR sample into males and females in sensitivity analysis, we identified an additional seven transcripts (three in males and four in females). None of these eight genes were replicated in the Finnish samples (which contained both men and women), although eight other transcripts located in these seven genes showed nominal significant association with zygosity in both cohorts. In the females-only analyses, but not in the males-only analyses, we confirm the finding of *PURG* indicating a potential sex-specific twinning maker.

The four extra genes identified in the females-only analyses all showed increased expression in the DZ twins and all show some relation to female reproduction. *ARSJ* and *UGT3A1* have a direct link to female fertility as both are involved in the synthesis of hormones and *ARSJ* is also associated with height in the FinnGen study (Mackenzie et al., 2008;Kurki et al., 2023)*. KIAAA0101* has been associated before with ovarian cancer and *KLK7* has been implicated with litter size in pigs (Dan & Yonggang, 2020; Jin et al., 2018; Kurki et al., 2023). Further studies are required to confirm a relation between these genes and twinning or female fertility, for example by investigating if these genes are differentially regulated in mothers of DZ twins around ovulation.

When comparing the current transcriptomics results with the TWAS results from the latest GWAS meta-analysis of DZ twinning (Mbarek et al., 2024), we find one overlapping gene; *PCBP2* was among the 5% transcripts with lowest p-values in the NTR results and showed lower expression in DZ twins. The TWAS analyses were performed in eight tissues mostly relevant to female fertility; breast, hypothalamus, ovary, pituitary gland, testis, uterus, vagina, and whole blood tissue, (Mbarek et al., 2024) while the current analysis only focused on RNA transcript levels in blood. In the TWAS, the whole blood data showed the lowest amount of genes with an association with DZ twinning, hinting that blood probably might not the most relevant tissue to study. Obtaining RNA transcript data from any other tissue at such a large scale is quite challenging. Additionally, the TWAS results predict gene expression based on genetic data, which cannot not take temporal fluctuations of RNA transcript levels within individuals into account (Gamazon et al., 2015). These natural fluctuations can be detected with transcriptomic data from blood samples and could explain why there is little overlap between the current study, and the previous TWAS.

We showed that the epigenetic signature of MZ twins is accompanied by a difference in transcript levels at two genes in blood: *HTT* and *FANCC*, are indicated both in the 5% transcripts with lowest p-values RNA transcripts and in the epigenetic MZ twinning signature (Bonder et al., 2017; van Dongen et al., 2021; Zhernakova et al., 2016). The former gene has been associated with Huntington’s diseases and the latter with a process that is activated when DNA replication is blocked called the Fanconi anemia pathway (Gordon & Buchwald, 2013; Jain & Roy, 2023). The increased methylation of these genes found by van Dongen et al. (2021) correlated with a lower expression of these genes in the MZ twins as shown in our study (van Dongen et al., 2021). For the remaining CpG sites in the epigenetic signature, we do not see any overlap with the transcriptomics results. This was not unexpected, since we previously showed that MZ-differentially methylated positions are strongly enriched in transcriptionally repressed, early embryonic expressed regions (van Dongen et al 2021).

We observe enrichment of the associated RNA transcripts in cell adhesion processes and the WNT-signaling pathways. Both were also enriched in the epigenetic signature and our results further underline the hypothesis that these processes may therefore be connected to the etiology of MZ twinning (Hernandez Gifford, 2015; van Dongen et al., 2021). The WNT signaling pathway or WNT signaling receptor activity was consistently enriched in all analyses. The SMAD and TGF-B pathways consistently showed enrichment in the FTC replication and NTR female-only analyses. *SMAD3* is the second top hit of the DZ twinning GWAS and also plays a central role in the TGF-B pathway as it is needed to translocate transcription factors to the cell nucleus and the TGF-B pathway has been implicated for DZ twinning before TGF-B/SMAD signaling is a well-established pathway involved in ovulation.

As for metabolites, we do not observe differences between MZ and DZ twins after correcting for multiple testing. We show that the three cohorts did not show strong correlations between coefficients, and even showed a negative correlation between the NTR and FTC, based on analyses in the total sample of males and females combined. The negative correlation could have been caused by menopause or contraceptive effects, which are known to influence metabolite levels (Bot et al., 2020; Hagenbeek et al., 2020) or the small sample of the FTC cohort. Furthermore, since dizygotic twinning depends on a double ovulation, a molecular signature of dizygotic twinning may disappear when ovulation stops beyond the female reproductive age or through the use of contraceptives. In the transcriptomics data, we performed a sensitivity analysis in the NTR female-only sample by removing all females that took contraceptives or indicated to be in menopause at the time of the biological sampling, and saw that the signal became stronger for genes in the SMAD pathway implicated in dizygotic twinning.

A final consideration of our study is that we did not have a non-twin control group. Therefore, for the transcriptome differences we observe between MZ and DZ twins, it is not necessarily evident whether they are connected to MZ or DZ twinning. However, some pathways clearly show overlap with previous DZ twinning GWAS results and thus are more likely driven by differential expression in DZ twins, whereas other transcriptomic results converge with the DNA methylation signature for MZ twinning (Mbarek et al., 2024; van Dongen et al., 2021).

Overall, we identified no metabolic differences and no replicating differences in gene expression levels of individual transcripts in blood between MZ and DZ twins. We do find limited replication of differentially expressed pathways in blood. Differential expression of TGF-B/SMAD pathway, involved in the regulation of ovulation and previously implicated in DZ twinning through GWAS emerges as an interesting potential transcriptional biomarker for DZ twinning and fertility that requires further confirmation in, for example, samples from mothers of DZ twins. More functional studies on twinning are to be encouraged, and pursuing efforts to include other omics (e.g. proteomics) and other tissues (e.g. ovary, and pre- and perinatal tissues) and other timing of transcriptomic and metabolomic measurements may pave the way to a better understanding of how these biological pathways are involved in twinning.

## Declarations

### Ethics approval

The study protocol was approved for the NTR by the Ethical Review Board of the VU University Medical Center and informed consent was obtained from all participants.

Ethical approval for all data collection waves from FT12 was obtained from the ethical committee of the Helsinki and Uusimaa University Hospital District and the Institutional Review Board of Indiana University. All data collection and sampling protocols were performed in compliance with the ethical guidelines. Parents provided consent for the twins aged 12 and 14 years old, while twins aged 17 and 22 years old provided written consent themselves for sample collection.

The study protocol for the FTC was approved by the Institutional Ethics Board of the Hospital District of Helsinki and Uusimaa, Finland (ID 154/13/03/00/11) and the Institutional Review Board of Augusta University.

### Availability of data and materials

The data of the Netherlands Twin Register (NTR) may be requested through the NTR data access committee (https://tweelingenregister.vu.nl/information_for_researchers/working-withntr-data).

The Finnish Twin Cohort data used in the analysis is deposited in the Biobank of the Finnish Institute for Health and Welfare (https://thl.fi/en/web/thl-biobank/forresearchers). It is available to researchers after written application and following the relevant Finnish legislation.

FinnTwin12 data analyzed in this study is not publicly available due to the restrictions of informed consent. Requests to access these datasets should be directed to the Institute for Molecular Medicine Finland (FIMM) Data Access Committee (DAC) for authorized researchers who have IRB/ethics approval and an institutionally approved study plan. To ensure the protection of privacy and compliance with national data protection legislation, a data use/transfer agreement is needed, the content and specific clauses of which will depend on the nature of the requested data.

All the codes used in this study have been uploaded to a publicly available GitHub page at https://github.com/hubersn1998/Omics_in_Twinning

### Authors contributions

NH, GD, RP and JvD have contributed to the conceptualization of the study. NH, GD, RJ and RP prepared the data for inclusion. NH has performed the analyses and was responsible for the first draft of the manuscript. RP, JvD, JJH, JK, GW and DIB provided supervision of the analyses and writing. GW and DIB were involved in the initial data collection of the NTR and MO and XW in the data collection of the FTC and FT12. All authors provided feedback and revision on the manuscript and supported the submitted version of the manuscript.

## Acknowledgement

This work largely builds upon the biobanking efforts performed by the BIOS and the BBMRI consortia and we thank all participants and staff for their contributions. Additionally we thank Dr. Fiona Hagenbeek for providing expertise and feedback throughout the project.

