## Supplement Figures for "Transcriptomic and Metabolomic analyses in Monozygotic and Dizygotic twins"

**Supplementary data**

Supplementary Figure 1: QQplot of the logistic regression performed in FTC in the top 5% RNA transcripts that showed the most differences in MZ and DZ twins in the NTR.

Supplementary Figure 2: Correlations between covariates in the FTC included in the RNA analyses.

Supplementary Figure 3: QQplot of 100 permutation of 1,325 random transcripts in the FTC. The red line indicates the qq-plot of the analyses of the 1,325 selected probes.

Supplement Figure 4: Enrichment of 1,325 random RNA probes on the FTC

Supplementary Figure 5: QQplot from the transcriptome analyses in the NTR including 42,663 RNA transcripts.

Supplementary Figure 6: Correlation between the effect sizes (betas) of the 1,325 top RNA transcripts in the NTR and the FTC cohort.

Supplementary Figure 7: Six most differentiated metabolites in the meta-analyses of the 196 metabolites.

Supplementary Figure 8: Correlation betas of the metabolites in the logistic regression in the three cohorts.

**Supplementary Figure 1**

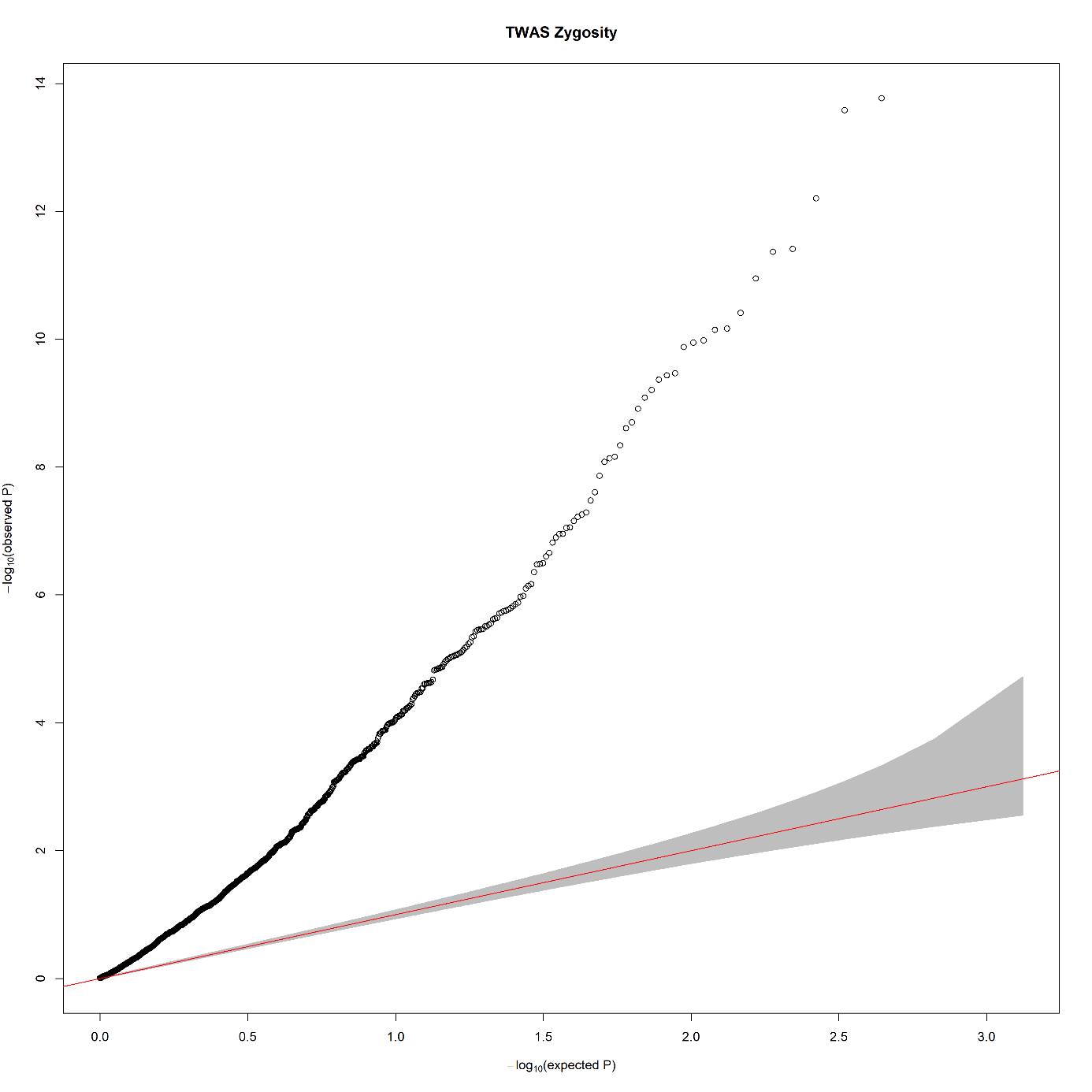

**Supplementary Figure 2**

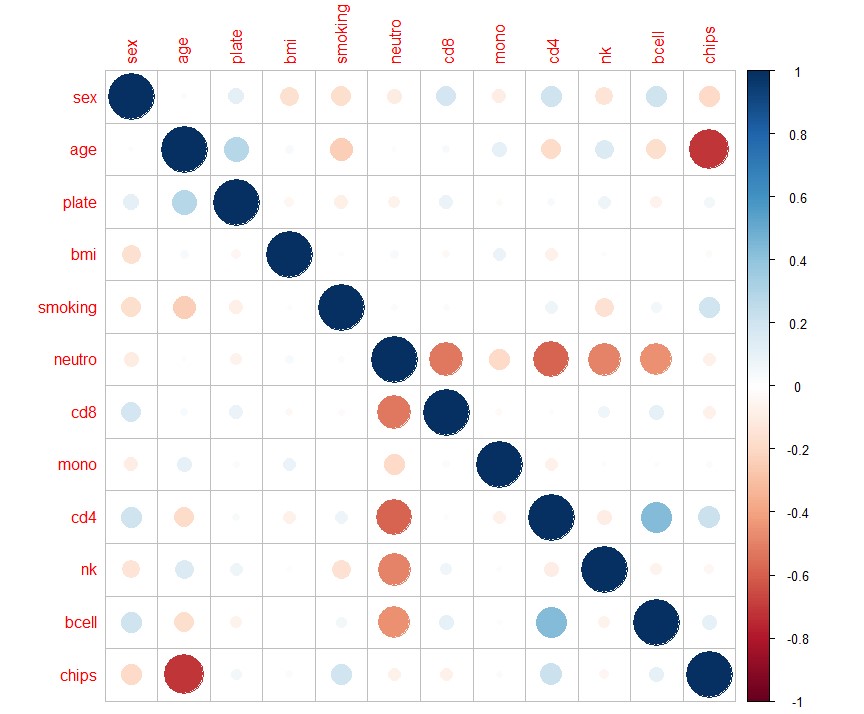

**Supplementary Figure 3**

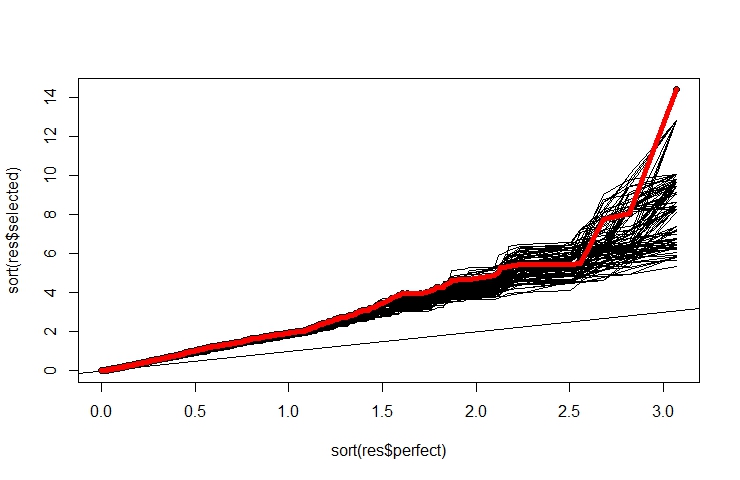

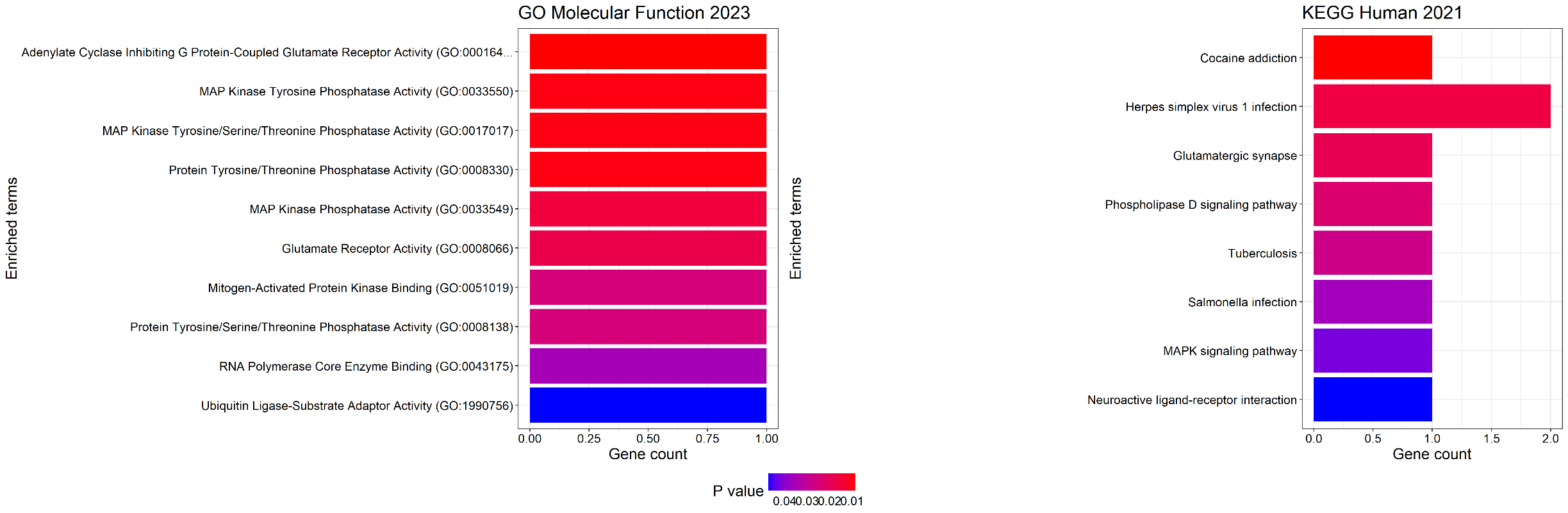
**Supplementary Figure 4**

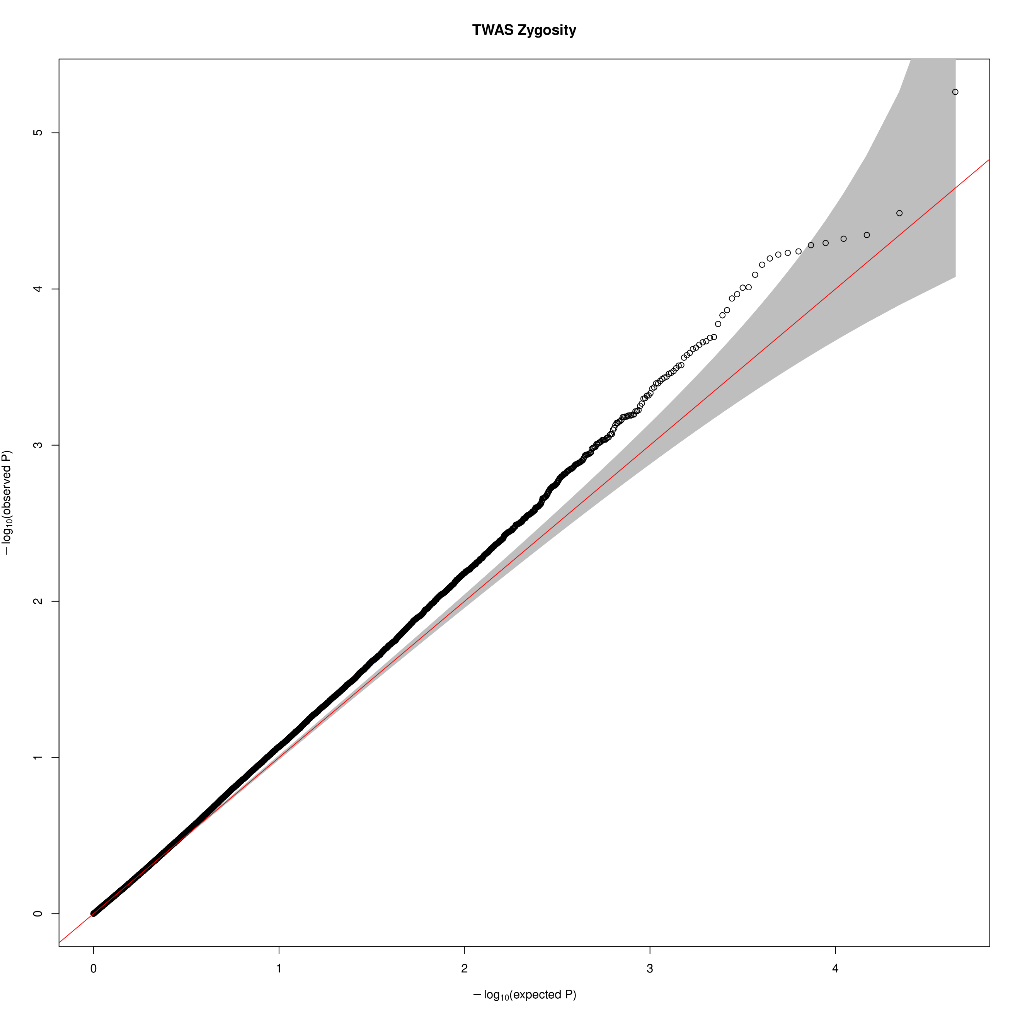
**Supplement figure 5**

**Supplementary Figure 6**
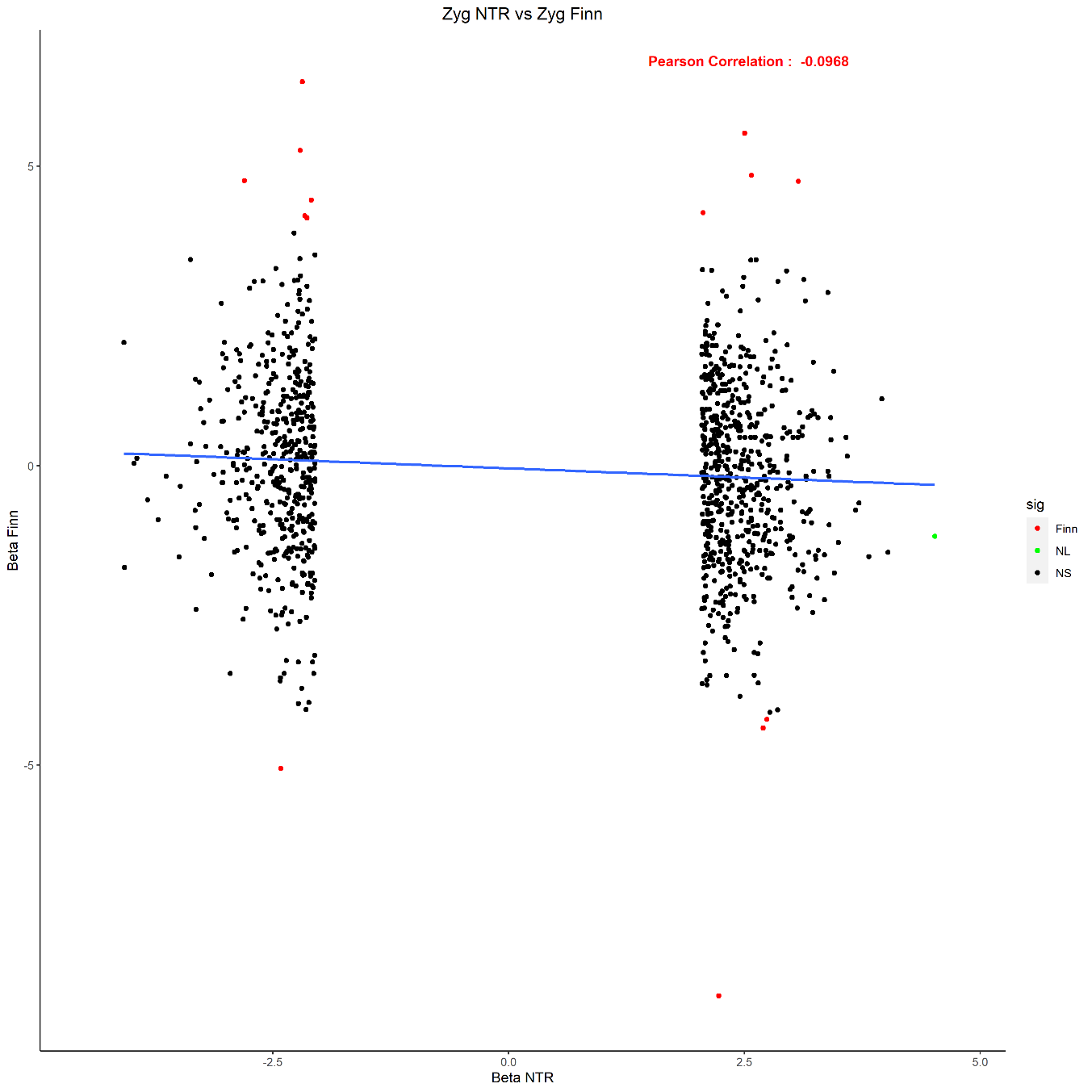

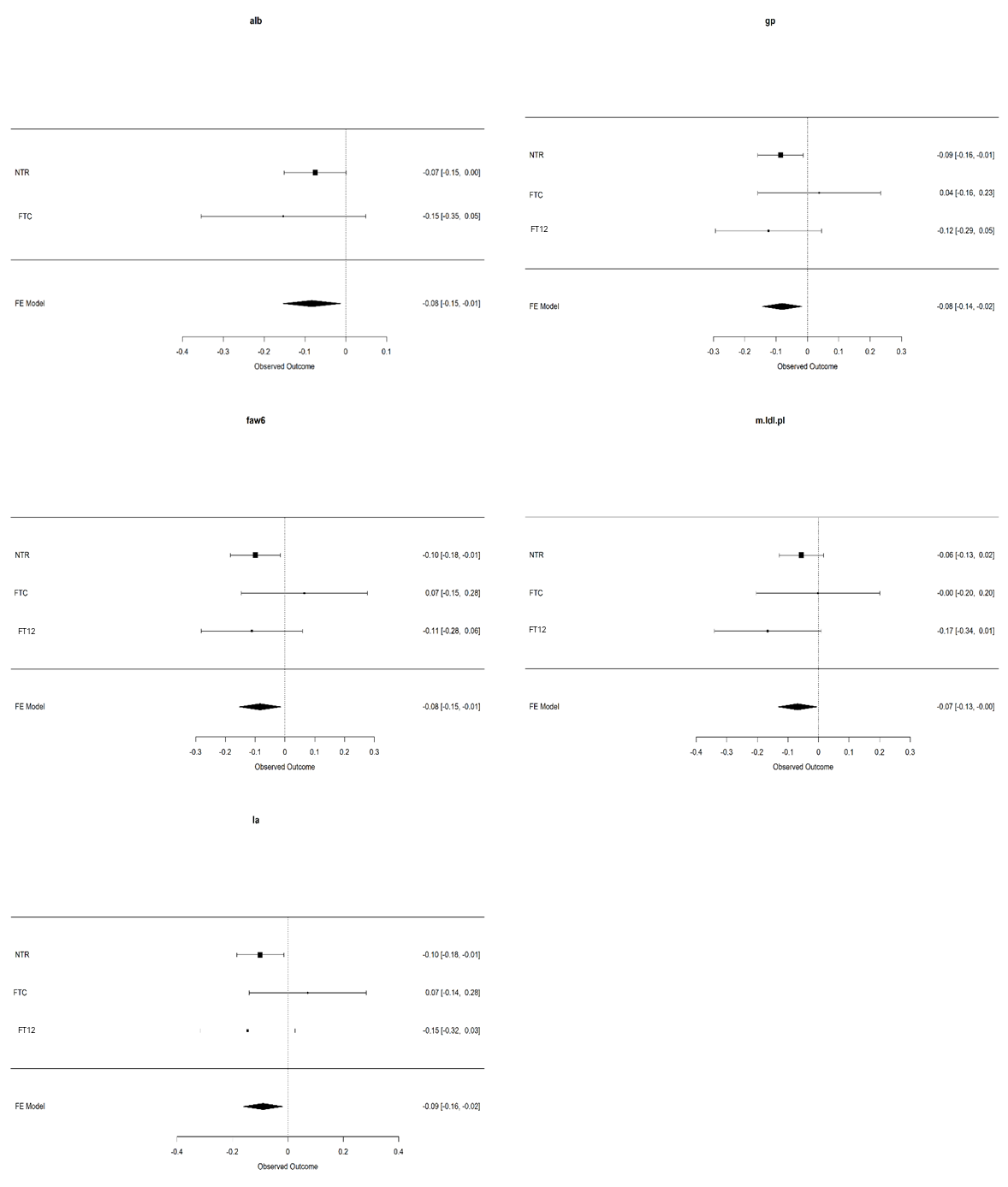
**Supplement Figure 7**

**Supplement Figure 8**

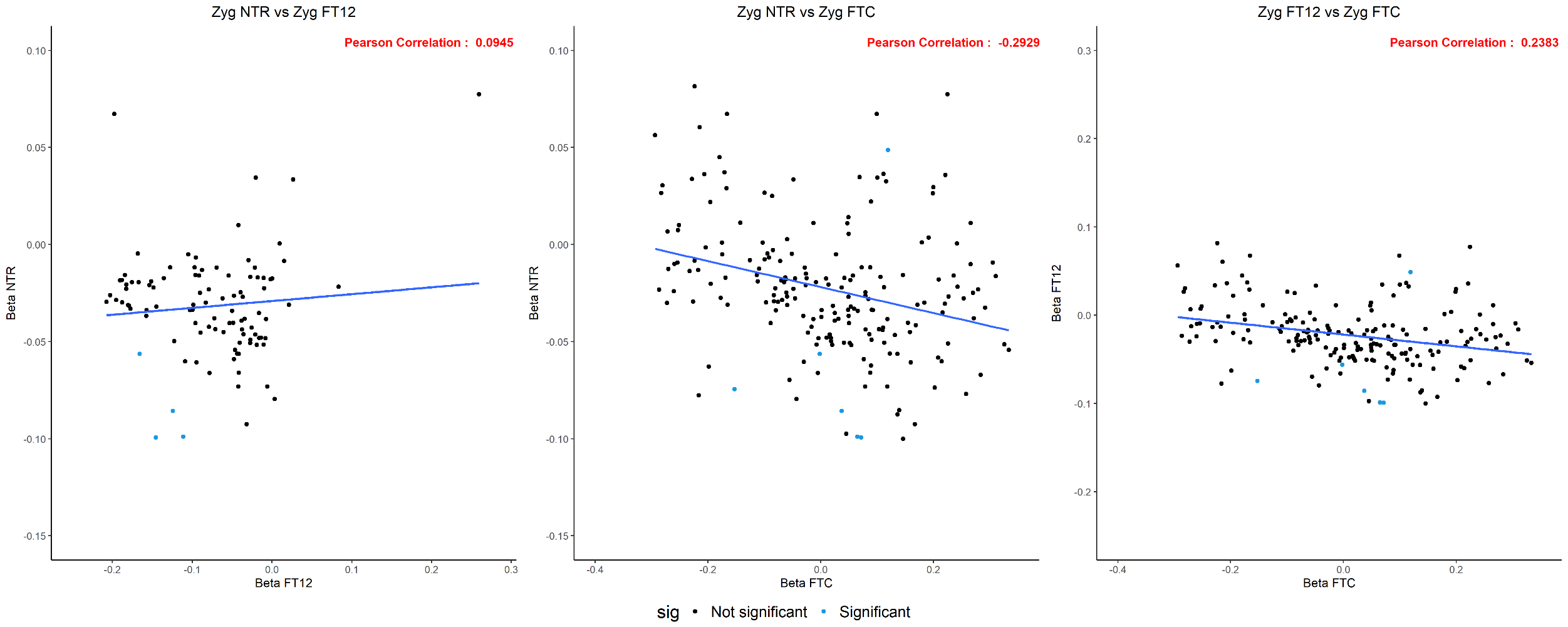
